# Use of Dry Blood Spots for Identifying Hepatitis E Infections

**DOI:** 10.1101/2025.11.09.25339677

**Authors:** Catia Alvarez, Robin Nesbitt, Kinya Vincent Asilaza, Pascale Sattonnet-Roche, Joseph Wamala, Shimul Das, Melat Haile, Monica Rull, Etienne Gignoux, Manuel Albela, John Rumunu, Isabella Eckerle, Iza Ciglenecki, Andrew S Azman, Benjamin Meyer

## Abstract

**Background:** Accurate and accessible diagnostics for hepatitis E virus (HEV) are essential for outbreak preparedness and surveillance, particularly in low-resource settings. Dried blood spots (DBS) offer a simple, scalable alternative to serum, but their diagnostic performance for HEV remains poorly characterised.

**Methods:** Paired DBS and serum samples were collected from suspected HEV cases during an outbreak in Bentiu, South Sudan. HEV RNA was detected and quantified by real-time RT-PCR, and anti-HEV IgM and IgG antibodies were measured by ELISA. DBS performance was compared to serum across assays, and associations between DBS positivity and serum viral load were analysed using logistic regression.

**Results:** Among 100 serum RT-PCR positive samples, 83 were positive by DBS (sensitivity 83.0%, 95% CI: 74.5-89.1). Ct values from DBS and serum were strongly correlated (ρ = 0.74), and most discordances at low viral loads (Ct > 30). DBS achieved 88.3% sensitivity (95% CI: 77.8-94.2) and 100% specificity (95% CI: 91.2-100) for IgM, while IgG sensitivity and specificity reached 95.0% (95% CI: 86.3-98.6) and 97.5-100%, respectively. Results were consistent across DBS card types and age groups.

**Conclusions:** DBS provide a reliable, practical alternative to serum for HEV diagnostics. Despite slightly reduced sensitivity at low viral loads, DBS maintained high specificity and strong correlation with serum results. Their ease of collection, storage, and transport without cold chain requirements supports their application for HEV surveillance and outbreak response in resource-limited settings. Further optimisation of elution methods to increase analyte concentration, along with improved storage conditions, could enhance diagnostic sensitivity.

## Introduction

Hepatitis E virus (HEV) infection is probably the most common cause of acute viral hepatitis and jaundice worldwide ^1,2^. It encompasses a single serotype and four human pathogenic genotypes. The prevalence of HEV infection varies between regions and depends on the genotype present. Genotypes 1 and 2 exclusively infect humans and are often associated with large-scale outbreaks and epidemics in low- and middle-income countries with low drinking water quality. The 2005 Global Burden of Hepatitis E study estimates 20.1 million annual infections, with 3.4 million clinical cases, 70,000 deaths, and 3,000 stillbirths attributed to HEV genotypes 1 and 2 ^3^. The true burden of HEV, however, remains unknown, mainly due to the absence of widely available diagnostics and the low specificity of suspected case definitions ^4,5^. Although the overall case fatality rate for HEV infection is around 1%, mortality can reach up to 25% among pregnant women in their third trimester ^6,7^. Studies from Bangladesh have reported that 19-25% of maternal deaths were associated with jaundice during pregnancy and suggested that HEV may be a causative factor, contributing to maternal mortality and adverse fetal outcomes, such as spontaneous abortion, stillbirths, and preterm delivery ^8,9^. In one study, 26.9% of HEV-positive pregnant women died, with fulminant hepatic failure identified as the primary cause of death ^10^. Together, these findings underscore the urgent need for improved diagnostics and surveillance to reduce the burden of HEV, particularly among vulnerable populations.

Current diagnostic approaches for confirming HEV primarily rely on serological assays such as enzyme-linked immunosorbent assays (ELISA) for IgM and IgG antibody detection or molecular techniques such as reverse transcription polymerase chain reaction (RT-PCR), typically performed on plasma or serum samples. These methods are essential for identifying acute infections, distinguishing past exposure, and confirming outbreaks. Their deployment in outbreak-prone regions, however, is often constrained by insufficient laboratory infrastructure, limited technical capacity, and the logistical challenges of collecting, storing, and transporting blood samples under strict cold chain conditions. These limitations highlight the urgent need for more flexible and accessible sampling methods to support both case detection and population-level surveillance.

Dried blood spots (DBS) represent a promising solution to these challenges. DBS are minimally invasive, can be collected with a simple finger prick, require limited training, and are more stable during storage and transport compared to venous blood, eliminating the need for a cold chain ^11^. Their utility has been demonstrated in the surveillance and diagnosis of multiple infectious diseases, including HIV ^12,13^, hepatitis B and C ^14,15^, and measles ^16,17^. Despite this, their application in HEV diagnostics remains underexplored. Most available studies on DBS and HEV focus on IgG antibody detection, with generally good concordance reported between DBS and serum samples. A validation study conducted in Bangladesh reported a sensitivity of 81% and a specificity of 97% for DBS-based IgG assays compared with serum ^18^. Similarly, a recent evaluation confirmed that DBS can reliably detect anti-HEV IgG, reinforcing their potential use in large-scale seroepidemiological investigations ^19^. Additional data from a study in pregnant women in Burkina Faso reported even higher performance, with DBS detecting anti-HEV IgG at 96.7% sensitivity and 100% specificity compared to serum ^20^. In contrast, evidence for IgM detection is more limited and less consistent. While some studies have shown promising accuracy with sensitivities of 86-91% and specificities up to 100% under controlled conditions ^21^, others have reported lower performance, including one study that found sensitivity and specificity of 76.7% and 93.8%, respectively, for DBS-based IgM detection ^20^. Similarly, DBS-based RNA detection by RT-PCR has shown good correlation with serum, but sensitivity is reduced in cases with low viral loads ^22^.

Improving HEV diagnostics is important for broader public health goals, including outbreak preparedness, transmission prevention, surveillance, and vaccine evaluation. Expanding the use of DBS could offer a practical and scalable solution for HEV diagnostics in low-resource settings. If DBS can be reliably used to identify both recent and past HEV infections, this would greatly expand surveillance capacity and simplify logistics for operational research and outbreak investigations.

To address these gaps, we investigated the performance of DBS compared to serum for both molecular and serological HEV diagnostics. We took advantage of paired serum and DBS samples collected from suspected hepatitis E cases during an outbreak in Bentiu, South Sudan, to understand the trade-offs in test performance. Specifically, we assessed detection of HEV RNA by RT-PCR and anti-HEV IgM and IgG antibodies by ELISA. By combining molecular and serological assessments, this study provides key evidence on the utility of DBS for HEV diagnostics in outbreak-prone, resource-limited settings.

## Methods

### Study setting

An enhanced hepatitis E surveillance system was set up to support operational research during the first emergency deployment of hepatitis E vaccine in the Bentiu Internally Displaced Person camp with approvals from the Médecins Sans Frontières (MSF) Ethics Review Board (ERB #2167) and by the South Sudan Ministry of Health Research Ethics Board (RERB-MOH #54/27/09/2022) ^23^. All individuals presenting to the Médecins Sans Frontières (MSF) hospital with suspected hepatitis E, defined as acute jaundice syndrome (AJS), characterised by the sudden or recent onset of yellow discoloration of the eyes or skin, often accompanied by dark urine and/or pale clay stools ^24^, were identified by clinicians and referred to the study team following their consultation or hospital admission. Study staff explained the objectives of the study to each suspected case. For those willing to participate, informed consent procedures were conducted: adults provided written informed consent, while individuals under 18 years of age provided written assent in conjunction with written consent from a parent or legal guardian. Upon consent, study staff administered a standardised questionnaire to collect information on symptoms, date of symptom onset, vaccination status, and sociodemographic details, including the participant’s address within the camp.

### Overview of Specimen Collection, Storage and Processing

A laboratory technician collected both venous blood and DBS and prepared all specimens for storage and transport. Venous blood was drawn, after which serum was separated by centrifugation at the MSF hospital. The samples were initially stored at the hospital in Bentiu and transported to Juba under -20°C conditions. Storage in Juba and shipment to Geneva was conducted at -80°C. For DBS, drops of venous blood were spotted directly onto filter paper cards, using either Whatman 903 Protein Saver Cards (n=40, Cytiva, Cat. No.10531018) or Ahlstrom Munksjö Biosample Collection Cards (n=131, Cat. No. 8.460.0012.A) and dried at room temperature. Once dried, each DBS card was placed in small plastic bags with a single desiccant to limit humidity exposure. All samples were then transported from the field to the Geneva Centre for Emerging Viral Diseases at the Geneva University Hospitals, Switzerland. Serum samples were shipped on dry ice with temperature loggers to ensure a -80°C cold chain during transport, while DBS were transported at room temperature. Upon arrival in Geneva, serum samples were stored at -80°C until testing, while DBS were kept at ambient temperature under dry conditions until analysis.

### Sample Selection

For the evaluation of RT-PCR, 100 paired DBS and serum samples were selected. To ensure representation across a range of viral loads, serum samples were stratified based on their cycle threshold (Ct) values into three groups: <25, 25-30, and >30. The study design aimed to include approximately one-third of the samples from each Ct range. However, due to the limited number of samples with Ct values <25, only 20 were available for inclusion in this group. Despite this slight imbalance, the final distribution was considered sufficient for robust comparison of DBS and serum performance.

For ELISA testing, 100 paired DBS and serum samples were selected, consisting of 60 ELISA-positive and 40 ELISA-negative serum samples. This distribution allowed for a more informative assessment of diagnostic sensitivity while still including a sufficient number of negative samples for specificity analysis. Selection was carried out in two steps: first, individuals who had already been tested by RT-PCR, ensuring that diagnostic results were available. As most of these individuals were seropositive for anti-HEV IgM and IgG, they provided the majority of positive cases. Additional negative samples were then included to reach the target of 100 pairs.

### Dried blood spots (DBS) preparation

For RNA extraction and RT-PCR, two spots from the DBS cards were punched using a hole puncher and placed into an Eppendorf tube containing 1 mL of EasyMAG lysis buffer (BioMérieux, Marcy-l’Étoile, France). Samples were incubated at room temperature for 30 minutes with slow vortexing to facilitate release of nucleic acids from the paper matrix. Following incubation, the lysates were centrifuged to remove residual filter paper debris. Based on spot input volumes (∼50-80 µL) and the typical recovery efficiency of ∼40-90%, each spot corresponds to approximately 20-70 µL of whole blood, equivalent to about 10-40 µL of serum ^25^. The clarified supernatant was then extracted and the RNA was quantified by RT-PCR. For comparison, RNA extraction from serum samples was performed using 200 µL of input material.

For ELISA, one spot from each DBS card was punched and placed in an Eppendorf tube containing 500 µL of elution buffer consisting of phosphate-buffered saline (Gibco PBS, Thermo Fisher Scientific) supplemented with 0.05% Tween-20 (PanReac AppliChem ITW Reagents) and 0.08% sodium azide (NaN_3_, Sigma-Aldrich) ^26^. Samples were incubated at room temperature on a laboratory shaker for 4 hours to ensure effective antibody extraction. Following incubation, eluates were centrifuged to remove paper debris, and the supernatants were used for ELISA. Considering that each DBS spot contained approximately 50-80 µL of whole blood and that antibody recovery efficiency typically ranges from 40% to 90%, this corresponds to about 20-70 µL of whole blood, equivalent to approximately 10-40 µL of serum after elution ^25^. For ELISA testing, 10 µL of the prediluted eluate was used per assay, while serum ELISAs were performed with 10 µL of serum.

### RNA extraction, reverse transcription polymerase chain reaction (RT-PCR) and viral load quantification

RNA was extracted from DBS samples using the NucliSens easyMAG instrument (BioMérieux, Marcy-l’Étoile, France), according to the manufacturer’s instructions. HEV RNA detection and quantification were performed using the ampliCube HEV 2.0 Quant real-time quantitative PCR (Mikrogen diagnostik, Neuried, Germany), which is designed for the detection and quantification of genomic RNA of HEV genotypes 1, 2, 3, and 4. The assay was carried out according to the manufacturer’s protocol. Viral load quantification was achieved by interpolating Ct values against a standard curve generated from quantified HEV RNA calibrators included in the kit. Results were expressed as international units per millilitre (IU/mL), taking into account the sample input and elution volumes: 200 µL for serum and 45 µL for DBS, with an elution volume of 50 µL for both. Samples were considered positive when the viral load was > 50 IU/mL. Amplification was performed on a CFX96 Thermal Cycler (Bio-Rad Laboratories, Hercules, USA), allowing simultaneous detection of HEV-specific RNA and the internal control within a single reaction.

### Enzyme-Linked Immunosorbent Assay (ELISA)

The Wantai HEV-IgM ELISA (WE-7196) and Wantai HEV-IgG ELISA (WE-7296) kits (Wantai BioPharm, Beijing, China) are commercial assays for the qualitative detection of anti-HEV antibodies. They use a two-step incubation, solid-phase capture ELISA in which recombinant HEV antigens coated on microwells bind specific IgM or IgG antibodies from the sample. Bound antibodies are detected with an enzyme-conjugated secondary antibody and visualised by a colorimetric reaction. For each sample, the absorbance value (S) is divided by the cut-off value (CO) to obtain the S/CO ratio. The cut-off value is calculated for each plate based on the mean absorbance of the negative controls and according to the manufacturer, S/CO ≥ 1.1 is positive, 0.9-1.1 borderline, and < 0.9 negative. Each kit includes positive and negative controls, and all reagents required for the assay.

### Statistical Analyses

Sensitivity and specificity were calculated with corresponding 95 % binomial confidence intervals (CIs) using the Wilson/Brown hybrid method as implemented in GraphPad Prism. To assess factors associated with DBS positivity, a multivariable logistic regression model was fitted in R (version 4.4). Explanatory variables included serum viral load, DBS card type (Whatman 903 or Ahlstrom Munksjö), and participant age group (<5, 5-14, and >15 years). All other analyses, as well as graphical visualisations, were performed in GraphPad Prism and Excel.

## Results

We selected 171 unique samples from suspected hepatitis E cases to compare diagnostic performance on DBS versus serum across three assays: RT-PCR, IgM ELISA and IgG ELISA (Table 1). The study population included 58 children under 5 years, 52 aged 5-14 years and 61 individuals over 15 years, with 74 (43.3%) females. The interval between sample collection and DBS testing ranged from 106 to 154 weeks.

**Table 1.** Description of the study population for RT-PCR and ELISA assays. Numbers represent paired DBS and serum samples collected from suspected HEV cases from Bentiu Internally Displaced Person camp, 2022. For RT-PCR, Ct categories were defined based on serum results.

| Assay | Total samples | Female (n) | Male (n) | Positives (n) | Negatives (n) | Age groups (n) | RT-PCR Ct categories (n) |
| --- | --- | --- | --- | --- | --- | --- | --- |
| RT-PCR<br>(Mikrogen<br>diagnostik) | 100 | 42 | 58 | 100 | 0 | 34 ($<5$ years),<br>34 (5-14 years),<br>32 ($>15$ years) | 20 (Ct $<25$),<br>43 (Ct 25-30),<br>37 (Ct $>30$) |
| IgM ELISA<br>WE-7196<br>(Wantai<br>BioPharm) | 100 | 47 | 53 | 60 | 40 | 23 ($<5$ years),<br>28 (5-14 years),<br>49 ($>15$ years) | - |
| IgG ELISA<br>WE-7296<br>(Wantai<br>BioPharm) | 100 | 43 | 57 | 60 | 40 | 46 ($<5$ years),<br>33 (5-14 years),<br>21 ($>15$ years) | - |

### RNA detection with PCR

For RT-PCR analyses, we selected DBS samples from 100 patients with serum samples confirmed positive by RT-PCR and obtained paired DBS specimens to capture a broad range of viral loads. The study population comprised 42 females and 58 males, with 34 children under 5 years, 34 aged 5-14 years, and 32 individuals older than 15 years. To assess performance across different levels of viremia, serum samples were stratified according to their Ct values into three groups: <25 (n=20), 25-30 (n=43), and >30 (n=37). Of the 100 serum samples positive by RT-PCR, 83 were also positive using paired DBS samples, corresponding to a sensitivity of 83.0% (95% CI: 74.5-89.1). Ct values obtained from DBS showed strong agreement with those from serum (Spearman ρ = 0.74), indicating good correlation between specimen types. Of the 17 DBS-negative samples, 15 had serum Ct values >30, suggesting reduced sensitivity of DBS at lower viral loads (Figure 1A). Logistic regression analysis indicated that for every one-log increase in serum viral load, the odds of DBS positivity increased 1.8-fold (95% 1.4-2.4), with no significant differences observed by DBS card type or age group.

**Figure 1:**
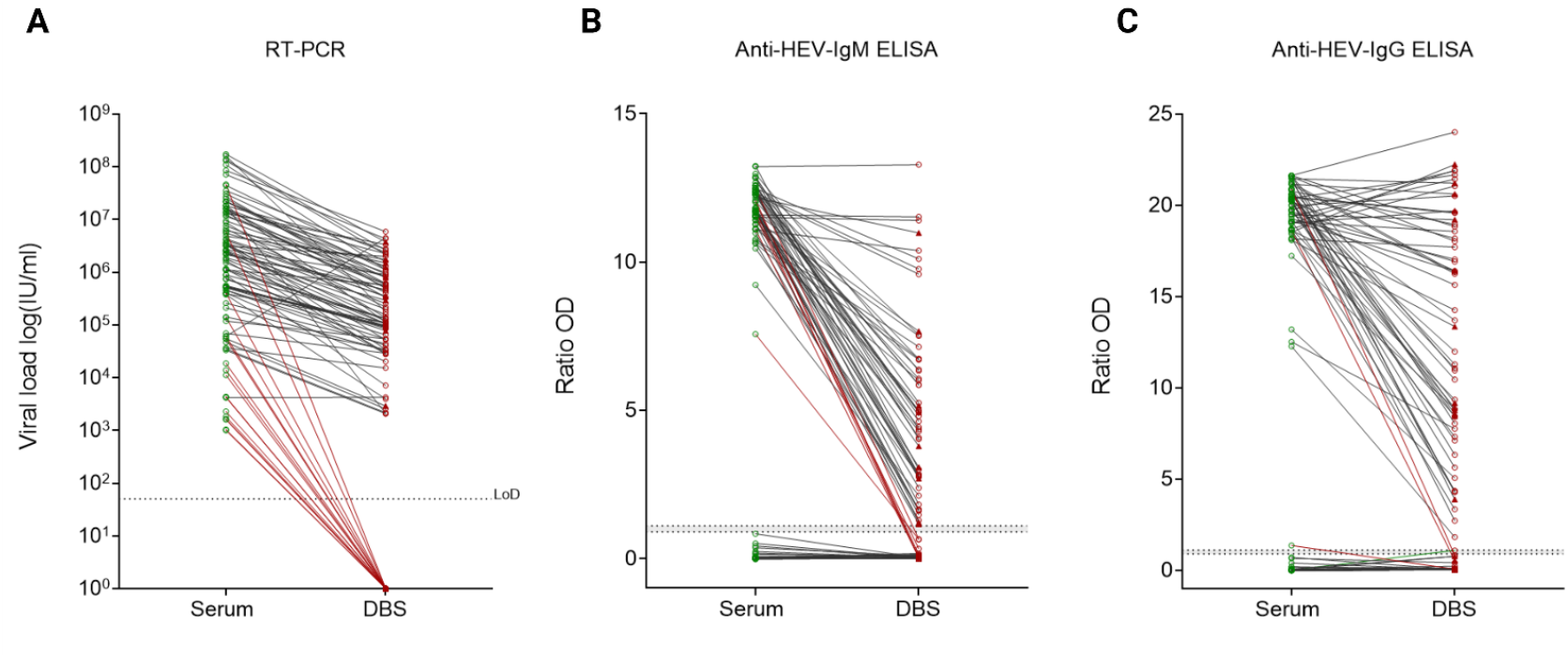
Comparison of DBS and paired serum samples for HEV. Results are shown for samples collected from suspected HEV cases during the Bentiu outbreak by (A) RT-PCR, (B) anti-HEV-IgM ELISA, and (C) anti-HEV-IgG ELISA. Serum results are shown in green and DBS results in red. The red line indicates samples that were positive in serum but turned negative in DBS assays, while the green line marks the serum-negative sample that was classified as borderline/indeterminate in the DBS assay. DBS samples collected on Ahlstrom Munksjö cards are represented by circles, whereas those collected on Whatman 903 cards are shown as triangles.

### Antibodies detection with ELISA

For ELISA testing, another set of 100 paired DBS and serum samples was selected, with 60 IgM-positive and 40 IgM-negative serum samples to allow a more robust evaluation of diagnostic sensitivity and specificity of DBS compared to serum. Among these, IgM analyses included 47 females and 53 males, distributed across three age groups: 23 children under 5 years, 28 aged 5-14 years and 49 individuals older than 15 years. For IgG analyses, 43 females and 57 males were included, with the following age distribution: 46 under 5 years, 33 aged 5-14 years and 21 over 15 years.

Of the 60 DBS samples that were serum IgM-positive by ELISA, 53 were also positive by DBS (sensitivity = 88.3%, 95% CI: 77.8-94.2) and all 40 serum IgM-negatives samples were correctly identified as negative by DBS, resulting in a specificity of 100% (95% CI: 91.2-100) (Figure 1B). For IgG, DBS correctly identified 57 of 60 serum IgG-positive samples, corresponding to a sensitivity of 95.0% (95% CI: 86.3-98.6). Among the 40 serum IgG-negative samples, 39 were negative and one was borderline/indeterminate, yielding a specificity of 97.5% (95% CI: 87.1-99.9) if the borderline is considered positive, or 100% (95% CI: 91.2-100) if considered negative (Figure 1C).

## Discussion

This study evaluated the performance of DBS for the detection of HEV RNA and anti-HEV antibodies in a population of suspected HEV cases during an outbreak in Bentiu, South Sudan. Using paired serum and DBS samples, which were stored for up to 35 months after collection, we observed that DBS-based RT-PCR detected HEV RNA in 83% of serum-positive cases, with strong correlation between Ct values from DBS and serum (Spearman ρ = 0.74). Discordant results for RT-PCR were primarily observed in samples with low viral loads (Ct >30), reflecting a reduction in sensitivity at the lower limit of detection. These findings are consistent with earlier work during a HEV outbreak in Darfur, where DBS and serum RT-PCR results showed 90.6% concordance ^22^.

For serological testing, our findings confirm that DBS-based ELISA can reliably detect HEV antibodies, though performance varies by antibody class. For IgG, DBS detected 57 of 60 serum-positive samples and 39 of 40 serum-negative samples, confirming excellent agreement for detecting past HEV exposure, with 95.0% sensitivity and 97.5% specificity. This accuracy is in line with prior studies, which reported sensitivities of 81-96.7% and specificities of 97-100% for DBS IgG ^18,20^. For recent or acute HEV infection, IgM detection from DBS identified 53 of 60 serum-positive samples and all 40 serum-negative samples (sensitivity 88.3%, specificity 100%), indicating slightly reduced sensitivity compared to serum but very high specificity. These results are consistent with previous reports ^21^. This reduction may reflect lower antibody concentrations in some samples, though we did not conduct quantitative ELISAs in this study. Together, these findings support the use of DBS for large-scale IgG seroprevalence studies and are feasible for IgM-based detection, with some sensitivity challenges if stored in similar conditions.

This study has several important limitations. The first relates to differences in sample volume between DBS and serum, which likely contributed to reduced sensitivity, particularly for RT-PCR and IgM detection. For RT-PCR, RNA was extracted from approximately 45 µL of serum derived from two DBS spots, whereas serum testing used 200 µL of input material. Similarly, for antibody detection, only one DBS spot (corresponding to ∼25 µL of serum diluted in buffer, of which 10 µL was applied to the assay) was used, compared with 10 µL of serum used in ELISA. These discrepancies in input volume and sample matrix could explain lower performance of DBS, particularly in specimens with low viral load or low antibody titers. Sensitivity could potentially be improved by optimising elution protocols, such as using smaller elution volumes to increase analyte concentration. Second, DBS samples in this study were prepared from venous blood rather than true capillary collection, which may limit comparability with field conditions where finger-prick sampling is typically used. Previous research in other infectious disease systems, including SARS-CoV-2 ^27^, HIV ^28^, and dengue ^29,30^, have generally shown that antibody and viral measurements obtained from capillary blood closely align with those from venous samples. However, minor discrepancies, particularly for IgM detection or at low viral concentrations have been reported. Third, DBS and serum samples were not tested simultaneously, and the storage duration and conditions for DBS were not optimal, raising the possibility that RNA degradation or antibody instability may have led to lower apparent sensitivity of DBS. Different storage conditions, such as maintaining DBS cards at 4°C instead of room temperature, and storage durations could help preserve RNA and antibody integrity over time. Concurrent testing of paired specimens would also minimise these effects and enable more accurate assessment of DBS performance without the effect of storage or time. Finally, the limited blood volume to DBS prevented precise quantification of viral load or antibody levels, restricting direct comparisons with serum and limiting interpretation of absolute concentrations.

Overall, our findings demonstrate that DBS from venous blood represent a feasible and practical alternative to serum for HEV diagnostics, particularly in field and outbreak settings. While DBS in this study were prepared from venous blood, the approach has the potential to be even less invasive if applied to capillary blood collected via finger prick. Their collection requires minimal training, and facilitates transport and storage without a strict cold chain, advantages that are especially valuable in resource-limited environments. By allowing detection of both recent and past infections, DBS can strengthen surveillance systems and facilitate operational research where conventional blood sampling is logistically challenging. While sensitivity is somewhat lower for samples with low viral load, and presumably with low-titer IgM, performance for IgG detection appears robust and IgM detection remains acceptable, supporting their application in both seroprevalence studies and outbreak investigations.

In conclusion, DBS from venous blood provide a reliable and efficient approach for HEV testing, offering high specificity and acceptable sensitivity in both molecular and serological assays. Their simple collection, storage, and transport procedures support their use for expanded surveillance, rapid outbreak response, and operational research in low-resource settings, while providing a foundation for future studies to optimise sampling methods, storage conditions, and assay performance.

## Data Availability

All data produced in the present work are contained in the manuscript

## Acknowledgements

We thank the MSF staff in Bentiu for data collection and the participants from the Bentiu IDP camp for their contribution. We also acknowledge the diagnostic laboratory of the University Hospitals of Geneva for assistance with serum serological testing.

